# Modeling the emergence of vaccine-resistant variants with Gaussian convolution COVID-19: Could the wrong strategy ruin vaccine efficiency?

**DOI:** 10.1101/2021.07.07.21259916

**Authors:** Christian Bongiorno, John Cagnol

**Author notes:** Equally contributing authors. Corresponding author: John Cagnol, CentraleSupélec, MICS, 3 rue Joliot-Curie, F-91192, Gif-sur-Yvette.

## Abstract

The SARS-CoV-2 virus, which is responsible for the COVID-19 pandemic, has been shown to mutate. In the absence of a vaccine, natural selection will favor variants with higher transmissibility rates. However, when a substantial portion of the population is vaccinated, natural selection will shift towards favoring variants that can resist the vaccine. These variants can therefore become dominant and even cancel out the benefit of the vaccine. This paper develops a compartmental model which simulates this phenomenon and shows how various vaccination strategies can lead to the emergence of vaccine-resistant variants.

## 1 Introduction

The COVID-19 pandemic is a crisis without precedent in modern history. The development of an efficient vaccine within less than a year is an equally unprecedented scientific and technological feat and provides a possible way out of the crisis. However, several challenges remain: the manufacturing and distribution of the vaccine, its societal acceptance, and the economic repercussions, to name but a few. The end of the crisis through vaccination also requires that variants of the virus will not develop a resistance.

The SARS-CoV-2 virus, which is responsible for the COVID-19 pandemic, is an RNA virus. These viruses tend to mutate at a higher rate than DNA viruses [4, 16], most likely due to the higher mutational frequency of RNA replicases compared to most viral DNA polymerases [5]. Therefore, this leads to swarms of genetic variants. Most mutations will lead to the death of the variant (lethal mutagenesis), some mutations will have no consequence on the virus’ ability to spread or to evade immunity, however, a few mutations may lead to more transmissible viruses or those that can resist the currently available vaccines [6].

The swarm of variants is subject to natural selection. In the absence of a vaccine, variants with higher transmissibility rates have an advantage against other variants with lower transmissibility rates. They will naturally tend to dominate. The “UK” B.1.1.7 variant, now denominated Alpha, was more contagious than the other variants, and thus quickly became dominant in early 2021 [8, 12, 13]. Before any vaccination attempts are made, vaccine-resistant variants do not have any advantage; if *V*_1_ is a vaccine-resistant variant but less transmissible than *V*_2_, which is not resistant to vaccines, then *V*_2_ will dominate over the *V*_1_ and the *V*_1_ variant may eventually go extinct. However, when a substantial part of the population has been vaccinated, the vaccine-resistant *V*_1_ may gain an advantage over *V*_2_ and thus take over.

This paper aims at investigating how various vaccination and non-pharmaceutical intervention (NPI) strategies, such as lockdown or other types of restrictions, can result in the emergence of vaccine-resistant variants.

Of course, such concerns are not new to the COVID-19 pandemic. Compartimental models have been used to study the effects of two competing strains of hepatitis B [26] and Mycobacterium tuberculosis [3]. More complex compartimental models have been used with three strains of the rotavirus in [22]. In the context of the common flu, [14] studied the emergence of transmissible resistant strains when using vaccine or anti-viral treatments. A comprehensive discussion of several attempts is available in [24] which reviewed 26 studied models of vaccine resistance for 12 different pathogens. However, what is new in our current context is the wide-spread scale of infection and the potential for a significant number of viruses, thus exacerbating the situation and even prolonging the pandemic.

In the context of COVID-19, the dynamics of two variants is investigated in [15, 17]. A control strategy is also proposed. The two-variant model is compared to real data in [18]. To the best of our knowledge, no work has been carried out on a generic large number of variants with different transmissibility and vaccine resistance characteristics.

Since the beginning of the pandemic, the impact of vaccine policies and non-pharmaceutical interventions has received considerable attention. One advantage to modeling is that many different simulations can be performed, thus allowing for the investigation of the dynamics of the pandemic under a wide variety of scenarii. The transmission dynamics and the thresholds of vaccine efficiency needed to prevent new waves of disease have been investigated in [25]. In [23], the authors use a sophisticated compartimental model (SIDARTHE) with ground data to simulate the outcome of several vaccination strategies. They find that non-pharmaceutical interventions (NPIs) have a higher effect on the epidemic evolution than vaccination alone, advocating to keep NPIs in place during the first phase of the vaccination campaign. The authors also mention there could be a risk of emergence of vaccine-resistant mutations, although their model does not take this into account. We refer to [9] for a more thorough review of the literature on the subject and to [20] for a thorough review on NPI.

There is growing literature on new variants of the SARS-CoV-2; see [8, 10, 12, 13] and references therein. However, it is not clear what further variants will occur and when. To the best of our knowledge, there is no model to predict such occurrences.

In Section 2, we propose a new model to take into account a large number of variants (essentially unlimited) with different transmissibility and resistance characteristics. The compartment of infected individuals is divided into sub-compartments to account for the various characteristics of the variants. To overcome the absence of a mutation model, we model the mutation using the multiplication by Toeplitz matrices representing a Gaussian convolution. This corresponds to giving a small probability of the virus to mutate with characteristics close to its own characteristics and a smaller probability to mutate to characteristics that are more remote from its current characteristics. In Section 3, we use our model to simulate three vaccine campaigns situation: vaccination in a community with a low level of infection, vaccination in a community with a high level of infection with and without NPI. Our model predicts that a vaccination campaign in a community with a low level of infection curbs the pandemic, however when the campaign occurs in a community with a high level of infection, the vaccination attempt fails if it is not accompanied with NPI. In Section 4, we investigate possible strategies of vaccination/NPI using the optimal control. We find that in some situations, nonintuitive strategies lead to a lower death toll. In Section 6, we present the limitations and caveats of our model.

## 2 The model

We consider an elementary compartimental epidemic model in the SIR family to describe the evolution of the COVID-19 pandemic, in a time period of a few months to a year, where:

- There is an effective vaccine for the initial strain
- There are variants with
  1. various transmission rates (linearly related to the reproduction number) and
  2. various resistance to the vaccine.

Variants will be classified according to criteria 1 and 2.

We consider *N* transmission rates, denoted *β*_1_ to *β*_*N*_, and *M* sensitivity rates to the vaccine, denoted *ω*_1_ to *ω*_*M*_. Subsequently, there are *N*×*M* classes. A variant with a transmission rate *β*_*i*_ and vaccine sensitivity *ω*_*j*_ will be called Variant (*i, j*). Note that two variants with the same transmission rate and the same sensitivity to the vaccine will be considered identical, which is a limitation of the model. Yet, a sufficiently large *N* and *M* resulting in a fine enough meshing should mitigate this impediment.

We shall consider the transmissibility rate to be increasing with the first index, meaning that if *i*_1_ *< i*_2_ then the variant with index *i*_1_*j* will be less contagious than the variant with index *i*_2_*j*. We shall also consider the resistance to the vaccine to be increasing with the second index, meaning that if *j*_1_ *< j*_2_ then the variant with index *ij*_1_ will be less resistant (more sensitive) to the vaccine than the variant with index *ij*_2_.

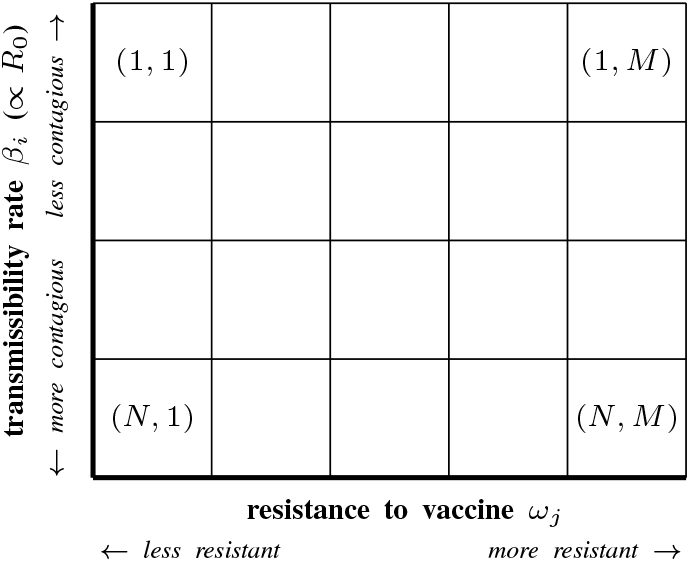

The population is divided into “compartments” which evolve as a function of time. The compartments are described hereafter.

### Compartments

#### Susceptible individuals

*S*(*t*) is the percentage of the population, which is susceptible to be infected, but has not been infected so far (up to time *t*).

#### Infectious individuals

Infected individuals are distributed in sub-compartments according to the variant they are infected with. Since the variants are numbered by a dual index (*i, j*) with *i* ∈ {1, …, *N*} and *j* ∈ {1, …, *M*}, we consider an array of compartments represented by an *N*×*M* matrix whose components *I*_*ij*_(*t*) describe the evolution of the percentage of the population infected with Variant (*i, j*). If no variant has transmissibility *β*_*i*_ and resistance *ω*_*j*_ then we have *I*_*ij*_(*t*) = 0 for all *t* ≥ 0.

#### Vaccinated individuals

V(t) is the percentage of the population which is vaccinated at time *t*. We make no distinction between the vaccines and assume vaccine-resistance is the same for all vaccines. Further development of the model should consider the variety of vaccines where variants could be more sensitive to one vaccine than another vaccine.

#### Recovered individuals

Recovered individuals are distributed in sub-compartments according to the variant the individual has recovered from. We consider the array of compartments represented by an *N*×*M* matrix whose components *R*_*ij*_(*t*) describe the evolution of the percentage of the population recovered with variant *ij* (*i* ∈ { 1, …, *N*} and *j* ∈ {1, …, *M*}. Our model accurately tracks an infection and a possible re-infection by another variant. The time frame of this work is a few months to a year. Thus, it is reasonable to neglect the declining acquired immunity and consider that re-infections by the *same* variant remain exceptional [11].

#### Deceased individuals

*D*(*t*) is the percentage of the initial population which, unfortunately, has died from the disease. We do not take into consideration births and only take into consideration deaths caused by the disease. Such extensions of the model could be made, but we do not believe they would have a significant impact on the results.

### Transition rates

#### Susceptible to Infected

Between *S* and *I*_*ij*_, the transition rate is assumed to be *S′* (*t*) = − *η*(*t*)*β*_*i*_*I*_*ij*_*S*. Following the standard SIR modeling assumptions, *β*_*i*_ *>* 0 is the average number of contacts per person per time, multiplied by the probability of the disease transmission in a contact between a susceptible individual and an individual infected with Variant (*i, j*). The function *η*(*t*) ∈ (0, 1] models non-pharmaceutical interventions such as stay-at-home orders. When *η* is equal to 1, no restriction is in place. When *η* is close to 0, the hardest restrictions are in place.

The rate *β*_*i*_ will be the discretization of the uniform distribution along 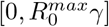 so the reproduction number is in 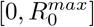. The coefficient *γ* is defined below. Thus, when *N >* 1, for any integer *i* in [1, *N*], we have

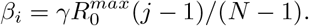

#### Susceptible to Vaccinated

We assume a rate of vaccination *n*(*t*).

#### Vaccinated to Infected

Between *S* and *I*_*ij*_, the transition rate is assumed to be *V ′*(*t*) =− *η*(*t*)*β*_*i*_*ω*_*j*_*I*_*ij*_*S* where *ω*_*j*_ measures the resistance of Variant (*i, j*) toward the vaccine. In other words, the efficiency of the vaccine is 1 − *ω*_*j*_; it is fully efficient when *ω*_*j*_ = 0 and useless when *ω*_*j*_ = 1. As before, *η*(*t*) models non-pharmaceutical interventions.

The rates *ω*_*j*_ will be the discretization of the uniform distribution along [0, 1]. Thus, when *M >* 1, for any integer *j* in [1, *M*], we have *ω*_*j*_ = (*j* − 1)*/*(*M* − 1).

#### Infected to Infected (emergence of variants)

We model the mutation by a transition from the infected compartment to the (same) infected compartment. This is performed using a discrete Gaussian convolution. This technique is widely used in image processing where it is known as a “filter”, notably to blur images. Details are provided in Appendix B.

One can define a fourth-order tensor *P* and compute the Gaussian convolution by computing the double contraction *P*.. *I*.

The standard deviation *σ* of the Gaussian convolution is the amount of “blurring” and represents the likelihood of (i) mutations to happen, (ii) leading to a viable virus, and (iii) with characteristics (*k, l*). The Gaussian distribution produces a small spill of the sub-compartments of *I* onto the sub-compartments that are nearby.

Because of the non-compact support of the Gaussian distribution, all sub-compartments will receive a positive value after the first convolution. Although this value is extremely small, the exponential nature of the dynamical system to come will amplify it. This is an unwanted situation that does not have any physical reality. Indeed, values in the compartments represent a fraction of the population. Any value under the inverse of the total population should be removed. Furthermore, differential equations models require the number of countable elements to be large enough to be represented by continuous variables. In other words, we should consider a tolerance level *h* below which, the spill-over is ignored. This value *h* should be at least larger than one over the population size. We will introduce a cut-off function *ψ* for compartment *I* with the property that *ψ*(*x*) = 0 when |*x*| *< h* and *ψ*(*x*) = *x* when |*x*| *>* 2*h*. To achieve the desired regularity on *ψ*, we define *ψ*(*x*) = − 3*x*^3^*/h*^2^ + 14*x*^2^*/h* − 19*x* + 8*h* for *x* ∈ [*h*, 2*h*] and *ψ*(*x*) = − 3*x*^3^*/h*^2^ − 14*x*^2^*/h*− 19*x*− 8*h* for *x* ∈ [− 2*h*, − *h*]. We shall denote Ψ the function from ℝ^*NM*^ to ℝ^*NM*^ which applies *ψ* to each component of the input matrix.

#### Infected to Deceased

Between *I*_*ij*_ and *D*, the transition rate is assumed to be proportional to the number of individuals infected with variant (*i, j*), which is the standard hypothesis in SIRD models. This rate is denoted *µ*. For the sake of simplicity, we consider all variants to have the same case fatality rate. Different case fatality rates could be considered by adding a dimension to matrix *I* (thus making it a third-order tensor) and adapting all depending tensors. This would complicate the model and we believe it would not drastically change the results unless there is a strong variation in this rate. Subsequently,

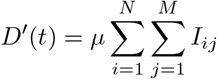

#### Infected to Recovered

Between *I*_*ij*_ and *R*_*ij*_, the transition rate is assumed to be proportional to the number of individuals infected with variant (*i, j*), that is 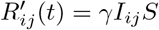. This is the standard hypothesis in SIR models. Again, we choose to consider that *γ* is constant across all variants. Adaptation of the model could be considered, but we believe it would not drastically change the results unless there is a strong variation in this rate.

#### Recovered to Infected

We consider a fourth-order tensor *ξ* = (*ξ*_*ijkl*_)_*ijkl*_ that indicates the transmission rate between the compartments *R*_*kl*_ and *I*_*ij*_. The components of this tensor represent the probability of disease transmission in a contact between an individual infected with the variant (*i, j*) and an individual who has recovered from the variant (*k, l*), multiplied by the average number of contacts per person per time unit. It is the counterpart of the coefficient *β*_*ij*_ for the susceptible to infected compartments.

The ratio 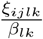 measures the consequence of the antigenic drift of the variant (*i, j*) towards a immunoresistant profile to the antibodies obtained after a recovery from variant (*k, l*). If it is equal to one, the prior infection by variant (*k, l*) provides no immunity toward the variant (*i, j*). If it is equal to 0, the prior infection by variant (*k, l*) provides no immunity to the variant (*i, j*).

As mentioned earlier, the time frame of this work allows one to neglect the declining acquired immunity and consider that re-infections by the *same* variant remain exceptional. Thus, we shall set

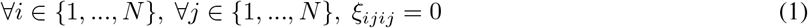

Identifying the remaining values of tensor *ξ* is delicate. The variants considered are theoretical and produced mathematically with the tensor *P*. They have not (yet) appeared in the real world. Subsequently, no experimental estimation of *ξ* can be performed. However, since the *j*-direction is used to model the impact of the antigenic drift on vaccine evasion, it is not unreasonable for 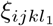 to be lower than 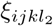 if *l*_1_ ≤ *l*_2_. We discretionally set *ξ*_*ijkl*_ to be 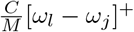, where *C* ∈ (0, 1) is a cross-variant re-infection constant. Consequently, if *l* ≤ *j* then for all *i* and *k* in {1, …, *N*}, we have *ξ*_*ijkl*_ = 0. This complies with (1) which states that a second reinfection by the variant is impossible. Furthermore, this choice of *ξ* implies that an individual infected by Variant (*i, j*) and then infected by Variant (*k, l*) (thus with *l > j*) cannot be reinfected by Variant (*i, j*).

### Summary of the model

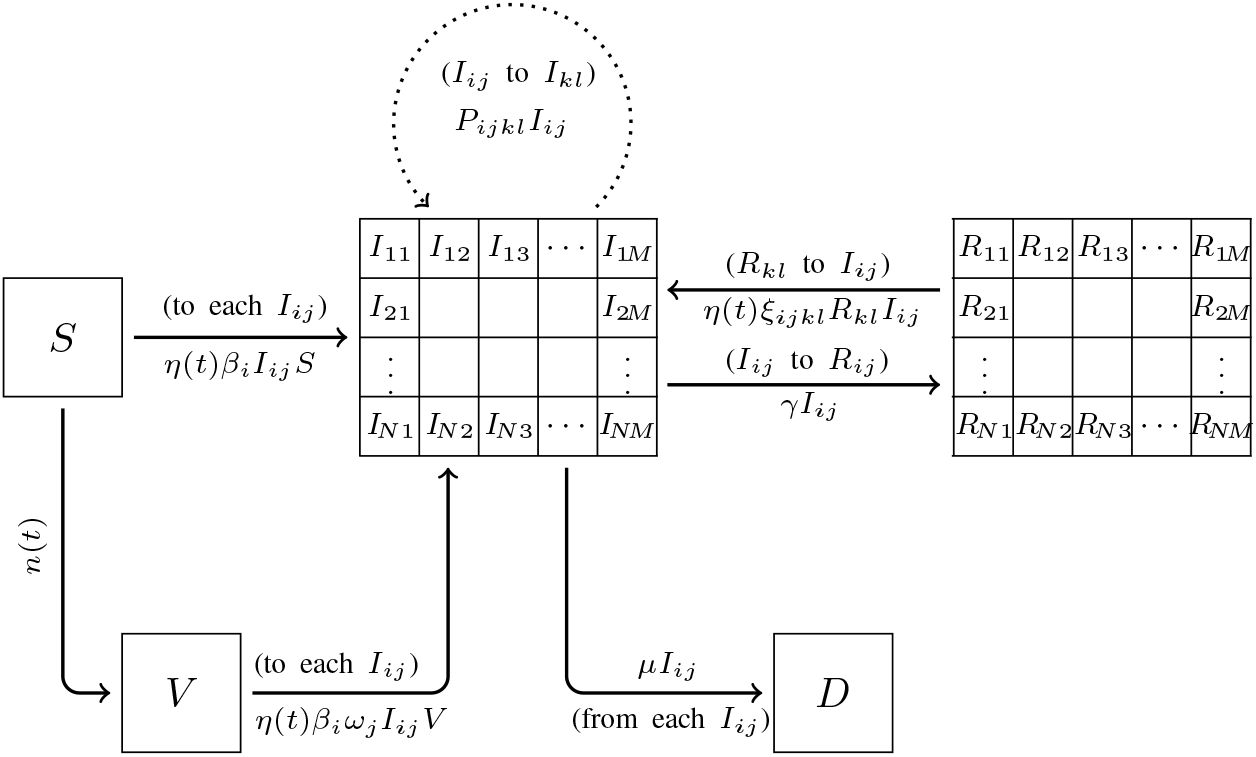

The model can be written in a more compact way using tensors. Tensors of order *n* are underlined *n* times, thus vectors are underlined once and matrices twice. The tensor product of vectors *u* and *v* is denoted *u*⊗*v* and is the matrix (*u*_*i*_*v*_*j*_)_*ij*_. The Hadamard product of two matrices *A* and *B* is denoted *A* ⊙ *B* and is the elementwise-product matrix (*A*_*ij*_*B*_*ij*_)_*ij*_. For a tensor *T*, the tensor *T*^*s*^ has indices which have been permuted according to permutation *s*. For instance, (*ξ*^(3,4,1,2)^)_*ijkl*_ = (*ξ*_*klij*_)_*ijkl*_. The vector <u>1</u> is the vector where all components are 1. Depending on the context, it can have *N* or *M* components. We refer to Appendix A for a more detailed presentation of these notations.

Because some of the compartments are matrices, the flux going out of these compartments will gather (sum up) when going into a scalar compartment. It is the case between compartments 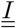 and *D*. Conversely, the flux going from a scalar compartment to a matrix compartment will split. It is the case between *S* and 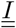 or *V* and 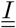 Finally, between two matrix compartments, the flux can braid. It is the case between compartments 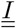 and 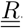.

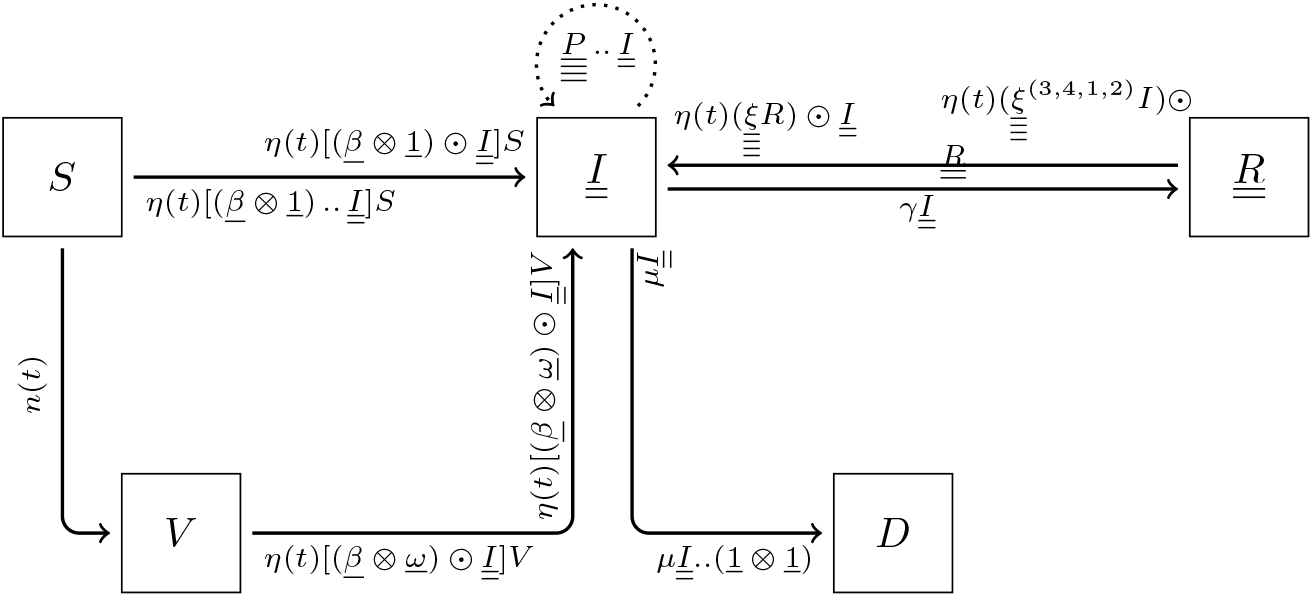

This leads to the differential system. Tensor underlining has been dropped for the sake of readability.

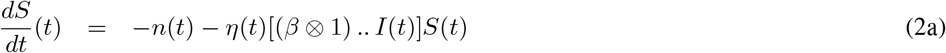

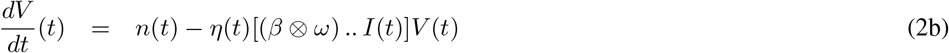

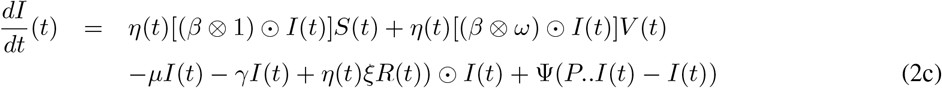

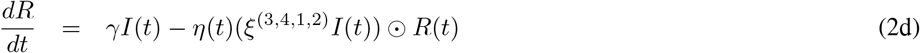

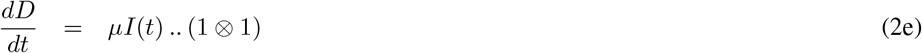

#### Proposition 1.

*System* (2) *has a unique solution on* [0, *T*].

*Proof*. Define *U* = [0, + ∞)× [0, + ∞)× [0, + ∞)^*NM*^× [0, + ∞)^*NM*^× [0, +∞) and consider the function *F* defined on [0, *T*] ×*U*→*U* such that 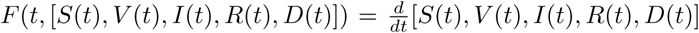. Function *F* is continuous with respect to *t* and Lipschitz-continuous with respect to the [*S, V, I, R, D*]. Existence and uniqueness of the solution to (2) is a straightforward consequence of the Picard-Lindelöf theorem. □

## 3 Numerical simulation of three scenarii

We simulate three vaccination campaign scenarii.

i. In a community with a relatively low level of infection (10 10^− 5^) with no NPI in place.
ii. In a community with a relatively high level of infection (200 10^− 5^) with no NPI in place.
iii. In a community with a relatively high level of infection (200 10^− 5^) and with NPI in place.

We consider a vaccination rate of 6 10^− 3^, which corresponds to a daily vaccination of 0.6% of the population. The vaccination campaign ends when the entire population has been vaccinated. We take *N* = 30 subdivisions of the transmissibility rate in the range 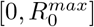 with 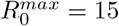. We take *M* = 40 subdivisions of vaccine efficiency. In all scenarii, we consider there is only one strain at time *t* = 0, which is completely sensitive to the vaccine. We set the recovery rate at *γ* = 0.09. We set the transmission rate to 0.2, which means that all components of the matrix *I*(0) are zero, but for the component (*i*_0_, *j*_0_) where 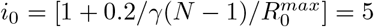 and *j*_0_ = 1. The component (*i*_0_, *j*_0_) of *I*(0) is set to 10 10^− 5^ and 200 10^− 5^ respectively for the communities with a low level and high level of infection. We consider that 25% of the population has been infected before and has recovered when the simulation begins at *t* = 0. We set the case fatality rate to *µ* = 2%.

The mutation parameter is set to *σ* = 1 and the reinfection parameter is *C* = 0.5. The population is one million people. These parameters are rather arbitrary and changing them will have an impact on the simulations. Therefore, the conclusions derived from these analysis are purely qualitative. Quantities are displayed to compare the situations but have no intrinsic meaning.

The simulation is performed on the time interval [0, 350] (days) using the explicit Euler method. The time step is 1. Figure 1 shows the evolution of the values in each compartment over time for scenarii (*i*), (*ii*), and (*iii*) and the distribution of variants.

**Figure 1.**
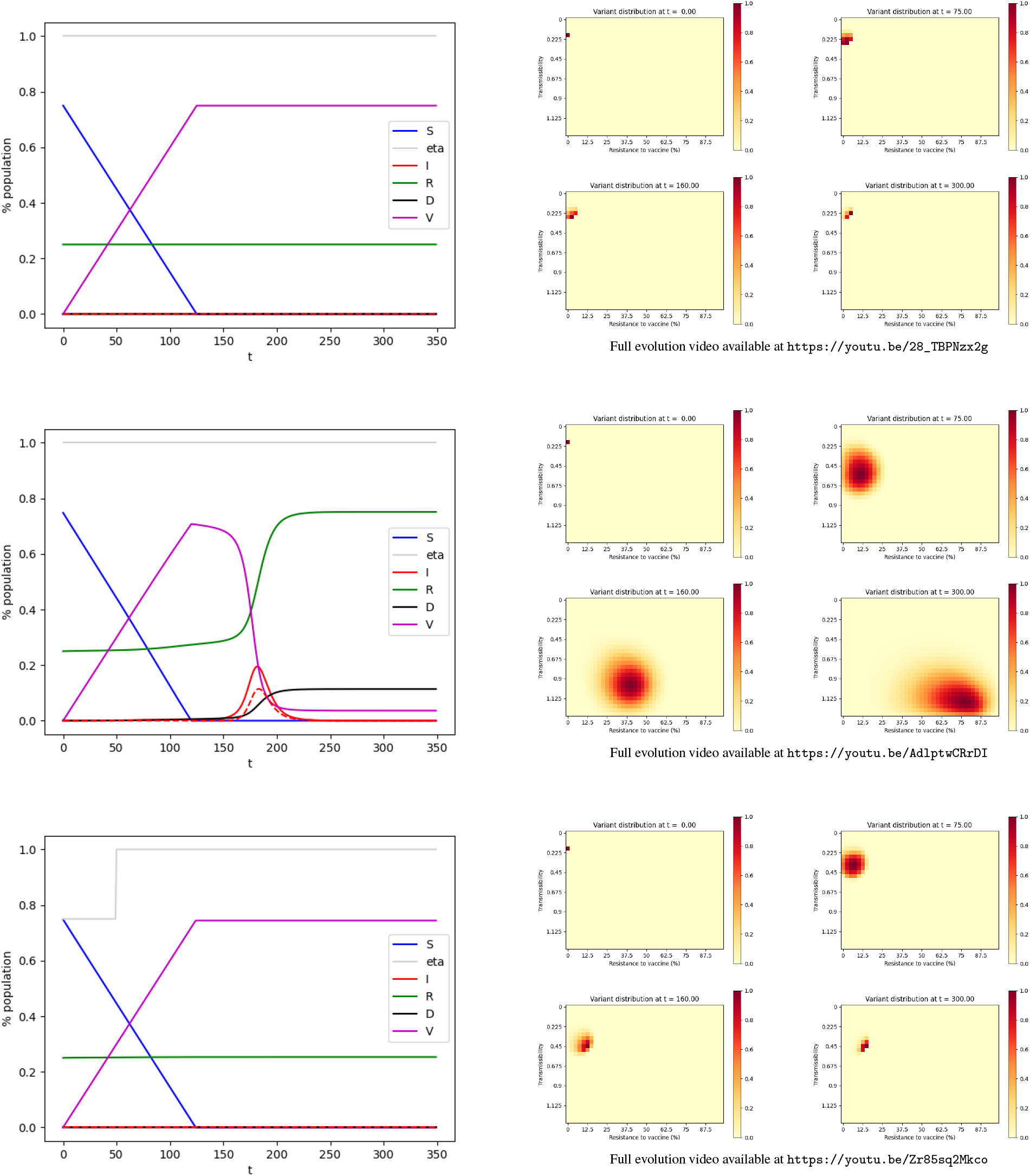
Evolution of the pandemic. Top: scenario (*i*) — Center: scenario (*ii*) — Bottom: scenario (*iii*) Left: Evolution of the compartments over time. Green: Susceptible individuals. Purple: Vaccinated individuals. Red: Infected individuals, the dashed line represents the individuals infected by variants resistant to a vaccine by more than 50%. Green: Recovered. Black: Deceased. — Right: Distribution of the variants at *t* = 0, *t* = 75, *t* = 160, *t* = 300.

In a community with a relatively low level of infection (10 10^− 5^) the vaccination campaign, with no NPI, curbs the pandemic. Other impending variants don’t have time to appear and the virus goes extinct. In the end (*t* = *T*), the death toll is roughly 0.004% of the initial population.

We now consider a community with a rather high level of infection (200 10^− 5^). In our model, the vaccination campaign with no NPI, fails to curb the pandemic. The vaccination gets rid of the initial strain, however, it also selects and advances variants that are resistant to the vaccine. As a consequence, a second peak of infection appears shortly after, with vaccine-resistant variants. It should be noted that the spike in infections by the variant occurs with a delay with respect to its emergence. In the end, the death toll is roughly 11% of the initial population.

We run our model for a community with a rather high level of infection (200 10^− 5^) where we put in place NPI on the interval [0, 50]. These restrictions are such that *η*(*t*) = 0.75 in this period. When NPI is put in place on this time interval, our model predicts the vaccination campaign curbs the pandemic. Other impending variants don’t have time to appear and the virus goes extinct. In the end (*t* = *T*), the death toll is 0.0006% of the initial population.

In conclusion, according to our model, the vaccination campaign shows desirable results if:

- It is carried out in a community with a low level of infections.
- It is carried out in a community with a high level of infection with NPI in place at least at the beginning of the campaign.

If the vaccination campaign is carried out in a community with a high level of infection, then nonpharmaceutical interventions are needed along with the vaccination campaign, at least at the beginning of the campaign, otherwise the vaccination campaign not only fails to curb the pandemic but it advances vaccine-resistant variants.

## 4 Optimal control

In this section, we investigate the optimal problem for the vaccination and NPI strategy. The natural and intuitive strategy would be to vaccinate as quickly as possible and to impose the strongest allowed NPIs. We aim to verify if is indeed the case.

In Subsection 4.1, we apply the Pontryagin principle to identify the location where the optimal control is located. We will see it is on the edges of a square. In Subsection 4.2, we identify the optimal control for the scenarii discussed in Section 3.

### 4.1 Identification of the location of the optimal control

Let us consider an observation period [0, *T*] where *T >* 0 is the final time. In the numerical simulations to come, we will chose *T* = 350. The time step will be set to 0.1 instead of 1 as in the previous section, for reasons that will be explained later.

Our goal is to minimize the number of deceased individuals on [0, *T*]. Thus, we shall consider the system cost

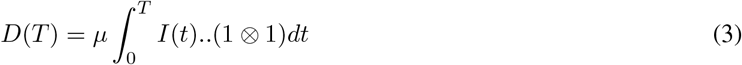

The control parameters are the vaccination rate *n*(*t*) and the non-pharmaceutical intervention *η*(*t*). These controls both come with constraints. The total number of vaccines used is constrained by the amount of vaccines available, the vaccination logistics, and possibly other factors. Let *n*_SUP_ represent this limitation:

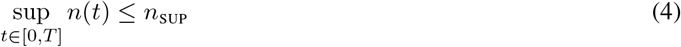

The non-pharmaceutical intervention *η*(*t*) is limited by socio-economic factors including the necessity to keep vital sectors running, social acceptability, and other factors.

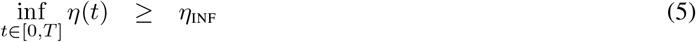

Let us consider the set of admissible controls:

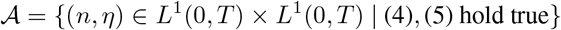

We use Pontryagin’s principle to address the optimal control problem. Consider a state function *t ⟼ x*(*t*), *t*∈ [0, *T*] with:

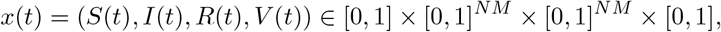

a control *u* ∈ 𝒜 and an adjoint-state *p ⟼ p*(*t*), *t* ∈ [0, *T*] (also known as costate) given by

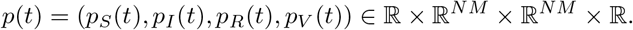

Let us define the Hamiltonian *H*:

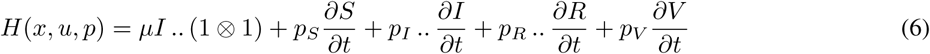

where 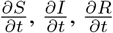 and 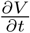 are given by (2).

Following the Pontryagin’s principle, if a control *t ⟼ u*^*^(*t*) is optimal, and *t ⟼ x*^*^(*t*) is the corresponding trajectory, it is necessary to have a nonzero adjoint vector function *p*^*^, which is the solution to

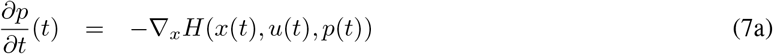

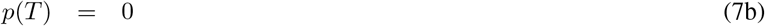

so that

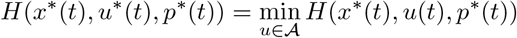

From (7) and some laborious computations, the adjoint variable *p* is the solution to:

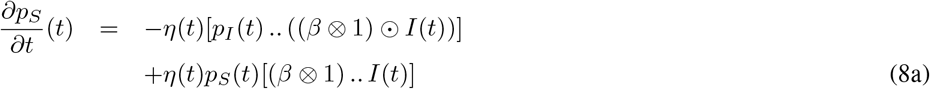

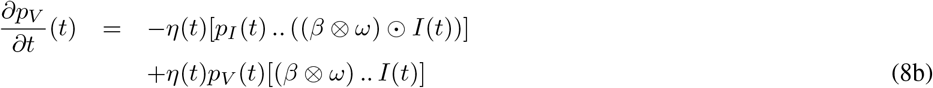

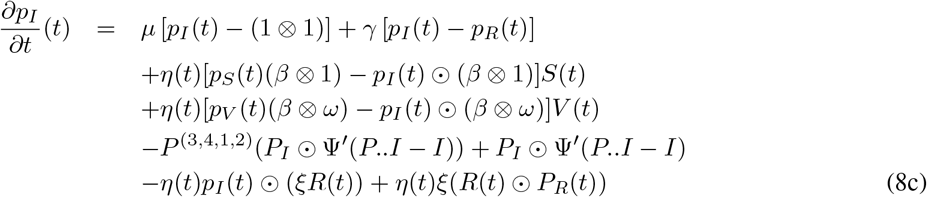

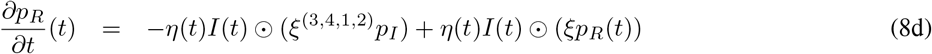

#### Remark 1.

*Here, the differentiation with respect to a matrix A is defined as the matrix with components that are the derivative with respect to the components of A*.

#### Proposition 2.

*There exists an optimal control u*^*^ ∈ 𝒜.

*Proof*. The existence of an optimal control is guaranteed by [7, Corollary 4.1] due to the linearity (thus, the convexity) of *µ I*.. (1⊗1) with respect to *u*, the a priori boundedness of the state solutions and the Lipschitz property of function *F* introduced in the proof of Proposition 1. □

To solve the minimization problem, we differentiate the Hamiltonian with respect to the control *u* = (*n, η*). The derivatives with respect to *n* and *η* are respectively

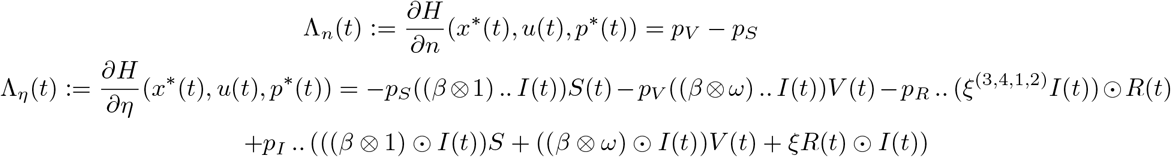

These non-vanishing terms suggest a bang-bang control, which is normal for such types of systems. Thus, the control *u* = (*n, η*) can be optimal only if it takes its values on the edges of the square as exposed in Figure 2. This is summarized in the following proposition.

**Figure 2.**
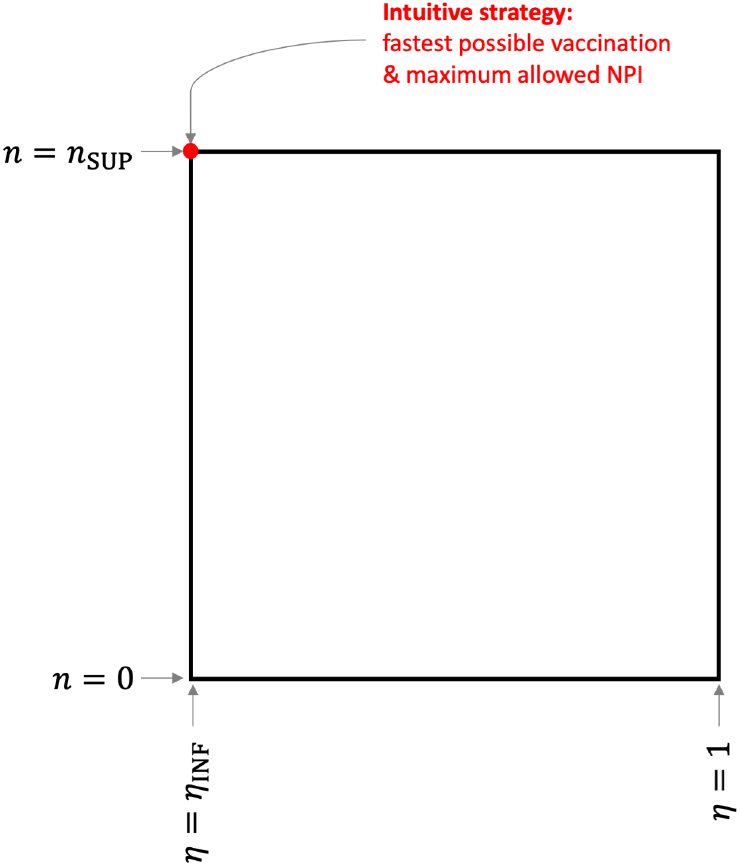
Control square: The optimal control *u* = (*n, η*) is located on the edges of the square.

#### Proposition 3.

*A necessary condition of optimality of the control u*^*^ ∈ 𝒜 *is that*

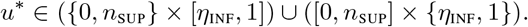

The north-west point of the square in Figure 2, represented in red, corresponds to the intuitive control which consists of vaccinating as quickly as possible and imposing the strongest NPI that is allowed:

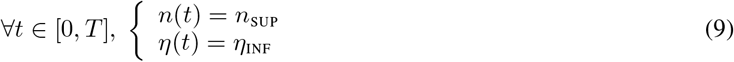

Nonetheless, there is no guarantee that this intuitive control is optimal. It is the case if Λ_*n*_(*t*) ≤ 0 and Λ_*η*_(*t*) ≥ 0 for all *t* ∈ [0, *T*]. However, if for some *t* we have Λ_*n*_(*t*) *>* 0 or Λ_*η*_(*t*) *<* 0, then the optimal control may not be the intuitive one. In this case, the optimal control will correspond to a point on the square, moving along the edges as time evolves, possibly stopping vaccination at times or lifting the restrictions.

### 4.2 Application to the three previously examined scenarii

Let us examine the situation of the community with a low level of infection with the parameters indicated in Section 3. We consider the fastest possible vaccination rate is *n*_SUP_ = 0.006 and we prohibit any NPI by setting *η*_INF_ = 1. In Section 3, we carried out the intuitive strategy (9), corresponding to the red dot on Figure 2, which consists of having *n*(*t*) = *n*_SUP_ until the compartment *S* is empty. The results were shown on the top part of Figure 1 and discussed earlier. By computing the adjoint state (8) and subsequently evaluating Λ_*n*_ and Λ_*η*_, we see that for all *t*∈ [0, *T*] we have Λ_*n*_(*t*) ≤ 0 and Λ_*η*_(*t*) ≥ 0 (see the top part of Figure 3). This indicates the intuitive control is indeed the optimal control.

**Figure 3.**
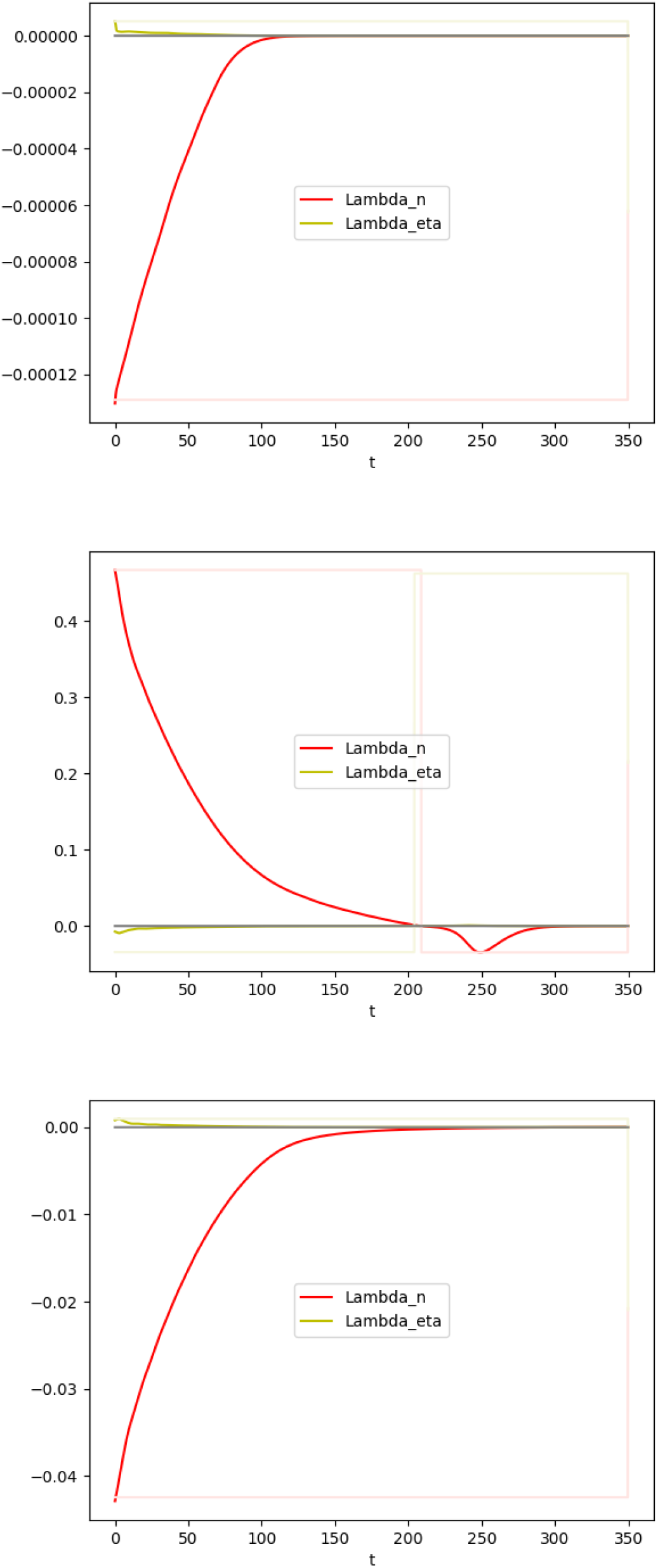
Adjoint state for the intuitive control strategy (9) in three scenarii. Red: Λ_*n*_. Yellow: Λ_*η*_. Top: scenario (*i*) — Center: scenario (*ii*) — Bottom: scenario (*iii*).

Let us now examine the situation of the community with a high level of infection with the parameters indicated in Section 3. Again, we consider the fastest possible vaccination rate is *n*_SUP_ = 0.006 and we prohibit any NPI by setting *η*_INF_ = 1. In Section 3, we carried out the intuitive strategy (9), corresponding to the red dot on Figure 2, which consists of having *n*(*t*) = *n*_SUP_ until the compartment *S* is empty. The results were shown on the center part of Figure 1. As discussed, this strategy does not curb the pandemic and the death toll is roughly 11% of the initial population. The question of the existence of a possibly better (non-intuitive) strategy remains. When computing Λ_*n*_ and Λ_*η*_ for the intuitive strategy, we obtain the values presented on the center part of Figure 3. The sign of Λ_*n*_(*t*) changes on [0, *T*]. This suggests the intuitive strategy is not optimal.

The optimal control for a community with a level of infection *and no NPI* is computed by two different methods. Due to the iterative process of both methods and the possible known unstability inherent to the explicit numerical scheme employed, we used a time step of 0.1.

**Method 1 — Computation of the optimal strategy using the adjoint state**. We start with *n* and *η* given by (9). We compute the adjoint state and we derive Λ_*n*_ and Λ_*η*_. When Λ_*n*_(*t*) ≥ 0 we set *ñ* (*t*) = 0, otherwise we set *ñ* (*t*) = *n*_SUP_. Similarly, when Λ_*η*_(*t*) ≤0 we set 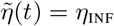, otherwise we set 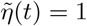. Then, we replace *n* by (1 − *λ*)*n* + *ñ* and *η* by 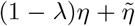, with *λ* = 1*/*10. The purpose of the coefficient *λ* is to avoid fast jumps that could result in a lack of convergence. Then we iterate this process until *n* and *η* no longer evolve from one iteration to the next (or for which the change is smaller than a tolerance threshold).

**Method 2 — Computation of the optimal strategy using the simulated annealing algorithm**. In Section 4.1, we established that the optimal control is located on the control square (see Figure 2). To find the close-to-optimal solution to minimize the system cost *D*(*T*), we discretize the square and address the optimization problem using a simulated annealing algorithm.

Both methods give comparable results. Method 2 relies on a coarser discretization to keep the dimension of the problem manageable, leading to slightly less precise results.

The strategy identified here is not intuitive; the vaccination campaign is delayed to prevent the emergence of the highly vaccine-resistant variants. This strategy results in a high death toll (9.7% of the population), but lower than the death toll resulting from the intuitive strategy consisting of vaccinating as quickly as possible with the highest possible NPIs (11%). We provide in Figure 4 the evolution of the pandemic when this non-intuitive strategy is used.

**Figure 4.**
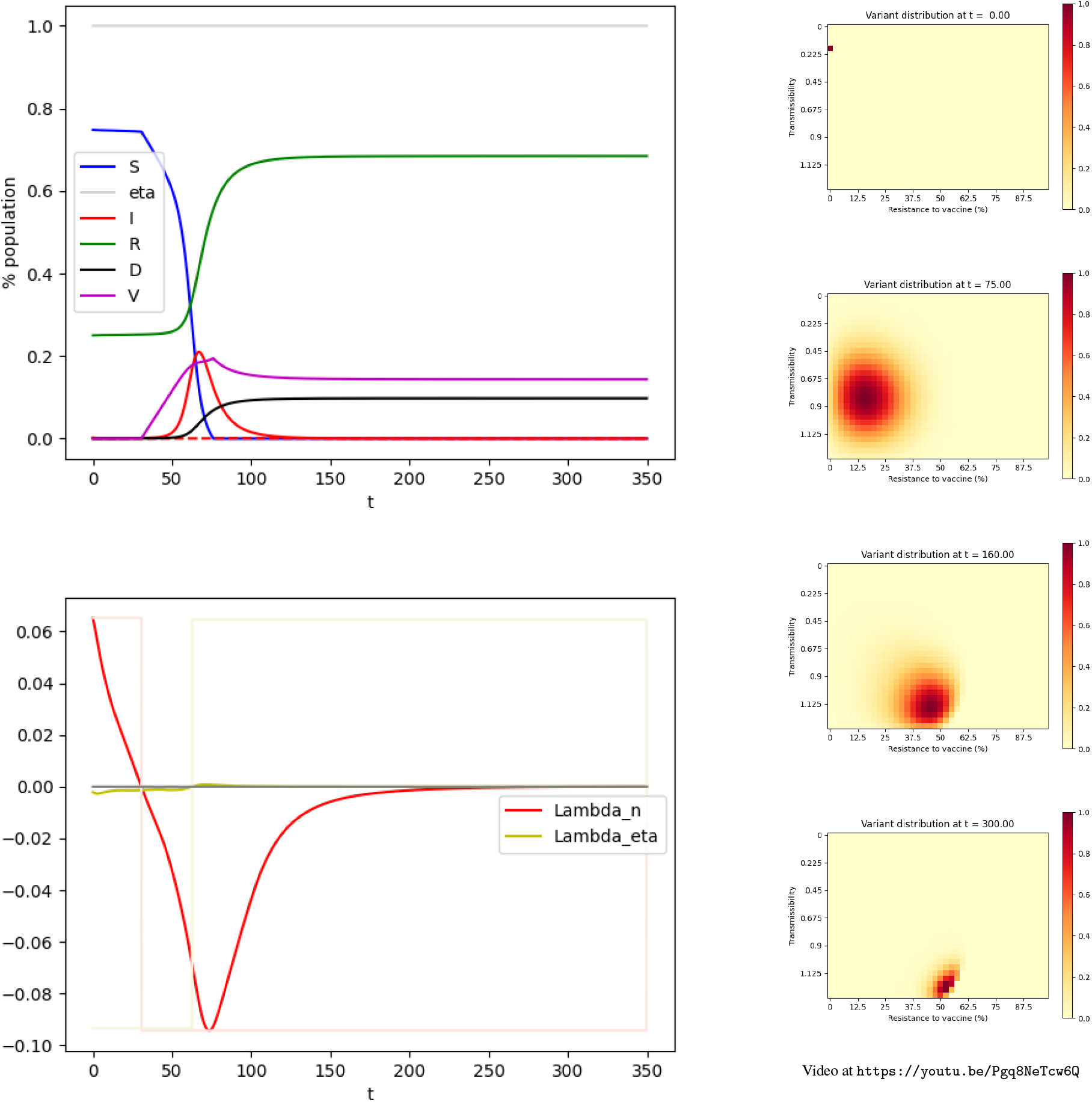
Optimal control for scenario (*ii*). Upper Left: Evolution of the compartments over time. Green: Susceptible individuals. Purple: Vaccinated individuals. Red: Infected individuals, the dashed line represents the individuals infected by variants resistant to a vaccine by more than 50%. Green: Recovered. Black: deceased. Lower Left: Adjoint state. Red: Λ_*n*_. Yellow: Λ_*η*_. Right: Distribution of the variants at *t* = 0, *t* = 75, *t* = 160, *t* = 300.

This result should not be interpreted as a suggestion to stop or delay vaccination campaigns in general. On the contrary, we support vaccinating and using NPI. When allowing NPI up to *η*_INF_ = 0.75 on [0, 50], the intuitive control (9) leads to Λ_*n*_(*t*) ≤ 0 and Λ_*η*_(*t*) ≥ 0 for all *t*∈ [0, *T*] as shown on the bottom part of Figure 3. This indicates the intuitive control is indeed the optimal control. In this case the death toll is 0.006% compared to 9.7% for the optimal vaccination strategy in the absence of NPI and 11% for the intuitive strategy in the absence of NPI.

## 5 Conclusion

In this paper, we have developed a compartmental model to account for several variants of SARS-Cov-2 with different transmissibility rates and resistance to a vaccine. We have used a Gaussian convolution to simulate the emergence of variants. Based on this modeling, we have run simulations for three different scenarii. While vaccinating is an effective strategy in a community with a low level of infection, and effective in a community with a high level of infection, if accompanied with non-pharmaceutical interventions (e.g. lockdowns), the model indicates that running a vaccination campaign in a community with a high level of infection while lifting all restrictive measures can lead to the selection of vaccine-resistant variants thus voiding the benefits of a vaccine.

In communities with a high level of infection, our model suggests that restrictions need to be kept in place during the vaccination campaign, or at least at the beginning of it. This goes against the understandable desire to go back to a “normal” life as quickly as possible, with the belief that vaccination is a way to avoid restrictions. Unfortunately, according to our model, lifting restrictions too soon may lead to selecting vaccine-resistant variants that could delay (and possibly jeopardize) the way out of the pandemic. This would become apparent with a delay and only when it is too late.

It should be emphasized that this paper does not recommend a vaccination strategy nor public health policy. It points out that, in some cases, vaccination may have serious adverse effects if not accompanied by non-pharmaceutical interventions. Clearly, further work is needed to better understand the virus mutation and adapt the mutation model of the *I*-compartment to a better fitted model. Hopefully, more precise mutation models can be achieved through ongoing and future research. The Biden administration is planning to spend $1.75 billion within Section 2402 of the “American Rescue Plan Act of 2021” [1] to strengthen and expand activities related to genomic sequencing, analytics, and disease surveillance [2].

## 6 Limitations and future work

As for any model, the one developed in this paper is an approximation of reality: simplifications are made, some phenomena are not taken into account, some parameters are estimated, or simply set to reasonable values. This model has shortcomings and is rather elementary. It focuses on (i) replacing the *I* compartment which customarily a scalar by a matrix and on (ii) using a Gaussian convolution to simulate the mutations.

We have built upon the SIR model, so the main ideas of the paper would be easier to understand. For better predictions, a more elaborate compartmental model would be better to take into account asymptomatic individuals, the age, and space structure of the population. The tensor-like model for SIR could be adapted to these more complex models at the cost of more variables to handle and possibly more tensors. We believe similar qualitative patterns to the ones found in this paper would be found with these more sophisticated models, but the values are likely to be affected.

We have not taken into account the waning immunity. This could be added to the model. Since it is not, the model developed here is not valid for long periods of time.

Due to the lack of robust models to describe the probability of emergence of variants with different characteristics, we have used a convolution mechanism to overcome this lack of data. Changing the values of *σ, M, N* will change the threshold values for the different behaviors, which, once again, calls for a qualitative interpretation of the results.

Finally, the model considers only one type of vaccine. If a vaccine-resistant variant emerges as computed in the simulation section of some scenarii and if new vaccines are manufactured to better target these variants, it will be useful to enrich the model to consider a panel of vaccines with their own efficiency with respect to each variant, and possibly the time necessary to manufacture these new vaccines. This would turn the scalar compartment *V* into a vector, vector *ω* into a matrix, and the real-valued function *n* into a vectorial function for which some components are set to zero until a new vaccine is manufactured.

## Supporting information

Fig 1 Video Scenario (i)

Fig 1 Video Scenario (ii)

Fig 1 Video Scenario (iii)

Fig 4 Video Scenario (ii) with optimal control

## Data Availability

Code is freely available.
https://github.com/cagnol/COVID-Variants

## Acknowledgments

Computations were performed using HPC resources from the Mesocentre computing facility at CentraleSupélec and École Normale Supérieure Paris-Saclay, which is partly funded by the CNRS and the Région Ile-de-France.

We would like to thank Professor Bernadette Miara, Professor Lulla Opatowski, Professor Romain Bourdais, Doctor Mohammad Ghousein, Professor Emmanuel Moulay and Professor Patrick Coirault for fruitful discussions. The second author would like to thank Bethany Cagnol who suggested he launch into this research.

## Appendix

### A Tensor notations

In order to obtain more concise notations, we use *tensors* to connect the compartments and describe their interactions. If some readers are not familiar with using tensors, they may simply consider them as multidimensional arrays. Such objects generalize vectors (first-order tensors) and matrices (second-order tensors). Roughly speaking, a third-order tensor is a stack of matrices. From here, we can continue to define higher-order tensors. Note that we are using a generalized definition of tensors so that each dimension does not need to have the same size, thus non-square matrices can be represented by a tensor.

The components of a tensor *A* of order *q* are denoted 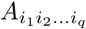.

Given two tensors *A* and *B* with the same order and the same size, one can define *A* + *B* as the tensor with this same order and size, whose components are simply the sum of the components of *A* and *B*.

Given two tensors *A* and *B* with the same order and size, one can define *A* ⊙ *B* as the tensor with this same order and size of *A* and *B* whose components are simply the product of the components of *A* and *B*. Such a component-wise product is called the *Hadamard product*.

Given two tensors *A* and *B*, respectively of orders *q* and *r*, the tensor product of *A* and *B*, which is denoted *A*⊗*B* is a tensor of order *q* + *r* whose components are 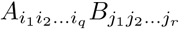. For example, if *A* and *B* are vectors (first-order tensors) of dimension *N* and *M*, then *A* ⊗ *B* is the *N* × *M* matrix

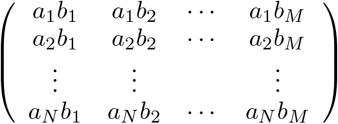

Subsequently, if we note 1 a vector where all entries are the real number one, then 1⊗ 1 is a matrix where all entries are the real number one.

Consider two tensors *A* and *B*, respectively of orders *q* and *r*, such that the last dimension of *A* and the first dimension of *B* are equal (say to *N*). We can define the simple contraction of *A* and *B*, and note it *A*.*B*, as the tensor of order *q* + *r* − 2 whose components are

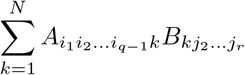

The simple contraction of a matrix (second-order tensor), with a vector (first-order tensor), is the usual product of the former by the latter. The simple contraction of two vectors (first-order tensors) is their usual inner product, thus a real number (zero-order tensor).

Similarly, given two tensors *A* and *B*, respectively, of orders *q* and *r*, such that the two last dimensions of *A* are equal to the first two dimensions of *B* (denoted to *N* and *M*), we can define the double contraction of *A* and *B*, and note it *A*..*B*, as the tensor of order *q* + *r* − 4 whose components are

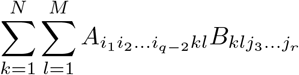

The double contraction of a fourth-order tensor and a matrix (second-order tensor) is a matrix (second-order tensor). The double contraction of two matrices (second-order tensors) is a scalar (zero-order tensor), in this case it is the Frobenius inner product of the matrices.

The transpose *A* of a matrix is unambiguously defined by permuting the indices. For a higher-order tensor, the permutation needs to be specified. For instance, given a fourth-order tensor *A*, one could define *A*^(3,4,1,2)^ as the tensor whose components are *A*_*klij*_, meaning the third index moves first, then the fourth, then the third, then the second. With this notation, the transposed of matrix *A* could be denoted *A*^(2,1)^.

### B Gaussian convolution

Let us consider an *N*×*M* matrix *I*. Our intent is to create a new matrix where each cell of *I* has “spilled” ever so slightly into the nearby cells.

Such operations are regularly performed in image processing through what is called a *Gaussian blur*. It is achieved through the convolution with the Gaussian function (*x, y*) *⟼ G*_*σ*_(*x*)*G*_*σ*_(*y*) where

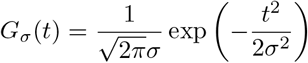

The parameter *σ* is the standard deviation of the Gaussian distribution and represents the amount of “spill-over” that is desired. The larger *σ* is, the stronger the blurring effect. In the context of this paper, this parameter *σ* models the probability the virus will mutate and leads to a variant with a different transmissibility rate or a different resistance to vaccine.

The value for the pixel (*i, j*) of the blurred image or, in the context of this paper, the proportion of infected individuals with Variant (*i, j*) is obtained by

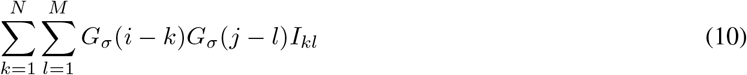

Note that in image processing, handling the edge is always an issue when performing a Gaussian blur. The problem is usually circumvented by modifying (10) to artificially extend the image by reflecting it with respect to the edge or replicating the last pixels. Less often, the problem is circumvented by taking all the values beyond the edge with the same constant. In the context of this paper, this latter solution is chosen with a constant equal to zero; it corresponds to the absence of spill-over from non-existing cells. Thus, expression (10) does not need to be changed.

Performing a Gaussian convolution of a vector of dimension *d* (signal stored in a 1D-array) can be achieved by multiplying the vector by a Toeplitz matrix 𝔗 (*d*) with elements

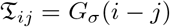

for *i* and *j* integers in [1, *d*] (see [19]). The extension of this method in the case of a matrix (or image stored in a 2D-array) does not lead to any difficulties.

Consider the fourth-order tensor

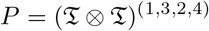

defined as the tensor product of 𝔗 (*N*) and 𝔗 (*M*), transposed by permutation (1, 3, 2, 4). This fourth-order tensor of dimension *N* × *M* × *N* × *M* can be written:

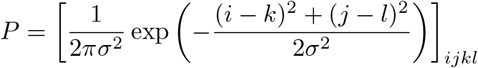

where *i* and *k* are integers in [1, *N*] and *j* and *l* are integers in [1, *M*].

Consider our *N* × *M* matrix *I*, we can compute the double contraction of *P* with *I*, which gives the matrix

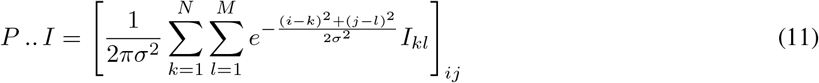

that we have represented in Figure 5.

**Figure 5.**
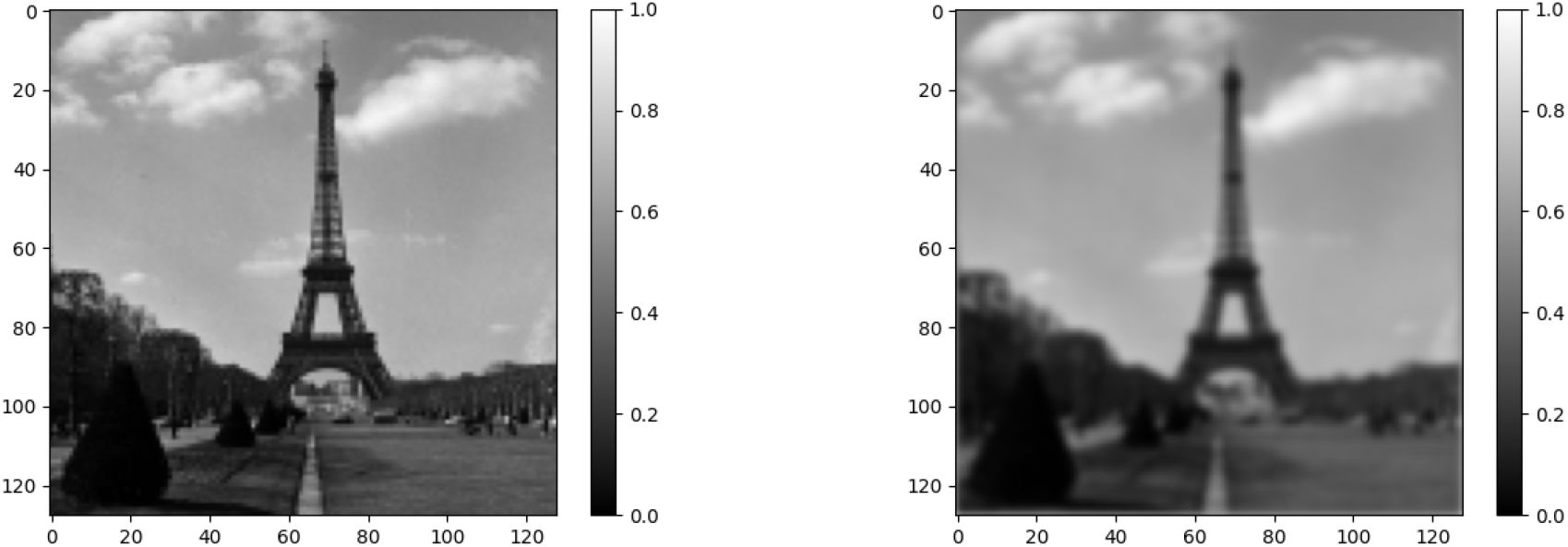
Gaussian Blur of the Eiffel Tower. The picture to the left is the original picture *I* (from a photo by Willem van de Poll released under CC0). It is a 128×128 matrix, the entries of which are a proportion of white. The picture to the right was obtained by computing *P*.. *I* as in (11).

While the use of Gaussian convolution is standard in image processing, the use of a fourth-order tensor to represent the operation is not. In the context of this paper, such a representation eases the computation of the differentiation of the Hamiltonian with respect to the state variables when applying the Pontryagin principle.

### C Model parameters

| Notation | Description | Range or Type | Notes |
| --- | --- | --- | --- |
| $N$ | Number of possible transmission rates in the model | $[1, +\infty)$ | (1) |
| $M$ | Number of possible sensitivity rates to the vaccine in the model | $[1, +\infty)$ | (1) |
| $T$ | Time observation period | $(0, +\infty)$ | |
| $t$ | Time | $[0, T)$ | |
| $S(t)$ | Proportion of healthy people (susceptible) at time $t$ | $[0, 1]$ | (2) |
| $V(t)$ | Proportion of vaccinated people at time $t$ | $[0, 1]$ | (2) |
| $I_{ij}(t)$ | Proportion of people infected by Variant $(i, j)$ at time $t$ | $[0, 1]$ | (2),(3),(4) |
| $I(t)$ | Matrix representing the proportion of infected people with respect to each variant at time $t$ | $N \times M$ matrix | (2) |
| $R_{ij}(t)$ | Proportion of people who recovered from Variant $(i, j)$ at time $t$ | $[0, 1]$ | (2),(3),(4) |
| $R(t)$ | Proportion of people who recovered at time $t$ | $[0, 1]$ | (2) |
| $D(t)$ | Proportion of deceased up to time $t$ | $[0, 1]$ | (2),(3),(4) |
| $\beta_i$ | Transmission rate from the susceptible to Infected in $I_{ij}$ | $[0, R_0^{max}]$ | (3),(8) |
| $R_0^{max}$ | Maximum basic reproduction number | $(0, +\infty)$ | |
| $\beta$ | Transmission rate vector | $[0, R_0^{max}]^N$ | (3),(8) |
| $\gamma$ | Recovery rate | $(0, +\infty)$ | |
| $\mu$ | Case fatality rate | $[0, 1]$ | |
| $\omega_j$ | Resistance of Variant $(i, j)$ to the vaccine | $[0, 1]$ | (9) |
| $\xi_{ijkl}$ | Reinfection rate to Variant $(i, j)$ for individuals who have recovered from Variant $(k, l)$ | $[0, +\infty)$ | (5),(6),(7) |
| $\xi$ | Reinfection tensor | 4th-order tensor<br>$N \times M \times N \times M$ | (4),(9) |
| $P$ | Mutation operator | 4th-order tensor<br>$N \times M \times N \times M$ | (11) |
| $\eta(t)$ | Non-pharmaceutical intervention coefficient at time $t$ | $(0, 1]$ | (10) |
| $\eta_{\text{SUSTAINABLE}}$ | Strongest possible restrictions sustainable over time | $(0, 1]$ | (10) |
| $\eta_{\text{INF}}$ | Strongest possible restrictions at a given time | $(0, 1]$ | (10) |
| $n_{\text{SUP}}$ | Maximum vaccination rate that can be achieved | $[0, +\infty)$ | |
| $n(t)$ | Vaccination rate at time $t$ | $[0, +\infty)$ | |

### D Scripts associated with this article

Three Python scripts are associated with this article:

- `simulation.py` approximates the dynamical system (2) with the Euler forward method and produces graphs regarding the evolution of the epidemic. It can also produce the distribution of the variants on the matrix. This script was used for Figure 1.
- `control-pontryagin.py` finds the optimal control (vaccination/NPI strategy) for a given situation. It relies on the computation of the adjoint state (8) and is described as Method 1 in Subsection 4.2. This script was used for Figures 3 and 4.
- `control-simanneal.py` finds the optimal control (vaccination/NPI strategy) for a given situation. This is Method 2 of Subsection 4.2. It relies on the simulated annealing algorithm and the Python module `simanneal` [21].

These scripts are released under the terms of the MIT license, are available at https://github.com/cagnol/COVID-Variants

